# Enhancing Drowning Prevention Efforts in Zambia: A Comprehensive Assessment

**DOI:** 10.1101/2025.09.08.25335300

**Authors:** Moses Mwale, Kelvin Mwangilwa, Sandra Musokota

## Abstract

**Aim:** This study examines the drowning prevention landscape in Zambia, identifying challenges, existing initiatives, and opportunities for improvement.

**Subject and Methods:** A convergent mixed methods approach integrated quantitative drowning case data from a WHO structured questionnaire with qualitative insights from structured and open ended responses. Data were collected in 2022 from 10 representatives across government, NGOs, community leadership, and public health expertise, consolidated via a stakeholder workshop. Quantitative data were summarized descriptively, while qualitative data were thematically analyzed to identify key themes, with findings integrated to assess convergence and divergence.

**Results:** Stakeholders identified males, children, youths, water related workers (e.g., fishers), and older adults as high risk groups, with inland water bodies (e.g., Zambezi River, Lake Bangweulu) as primary drowning locations during fishing, transportation, and water collection. Zambia Police reported 89 drowning deaths in 2022 (88.8% male, n=79; 11.2% female, n=10), primarily among individuals aged 30–49 years (n=50). Key barriers to drowning prevention include the absence of a national coordination mechanism, limited implementation of interventions (e.g., swim education, physical barriers), weak enforcement of boating safety laws, and fragmented data systems. National strategies lack specific targets, and no legislation mandates lifeguards or pool fencing.

**Conclusion:** A coordinated, multi sectoral approach involving capacity development, stakeholder partnerships, advocacy, and international support is critical to address these gaps and reduce drowning mortality in Zambia.

## Introduction

Drowning is a significant global public health challenge, causing an estimated 236,000 annual deaths and ranking as a leading cause of injury-related mortality, particularly among children and young adults in low- and middle-income countries (LMICs) (Meddings, Scarr, Larson, Vaughan, & Krug, 2021; World Health Organization, 2014; Franklin et al., 2020). In Africa, drowning is a substantial contributor to injury-related deaths, especially in rural areas where access to safe water sources is limited (Peden, Oyegbite, Ozanne-Smith, Hyder, Branche, Rahman, Rivara, & Bartolomeos, 2008; Bowman, Seedat, Duncan, & Kobusingye, 2006). Zambia, a landlocked Sub-Saharan African nation with a population of approximately 19 million (Zambia Statistics Agency, 2022), faces a considerable drowning burden, with WHO Global Health Estimates reporting approximately 600 drowning deaths in 2019 (5.8 per 100,000) (World Health Organization, 2019). Figure 1 illustrates the trend of drowning deaths in Zambia from 2000 to 2019, highlighting a persistent burden (World Health Organization, 2019). Local data likely underreport this burden due to fragmented surveillance systems, underscoring drowning’s profound impact on national health.

**Figure 1:**
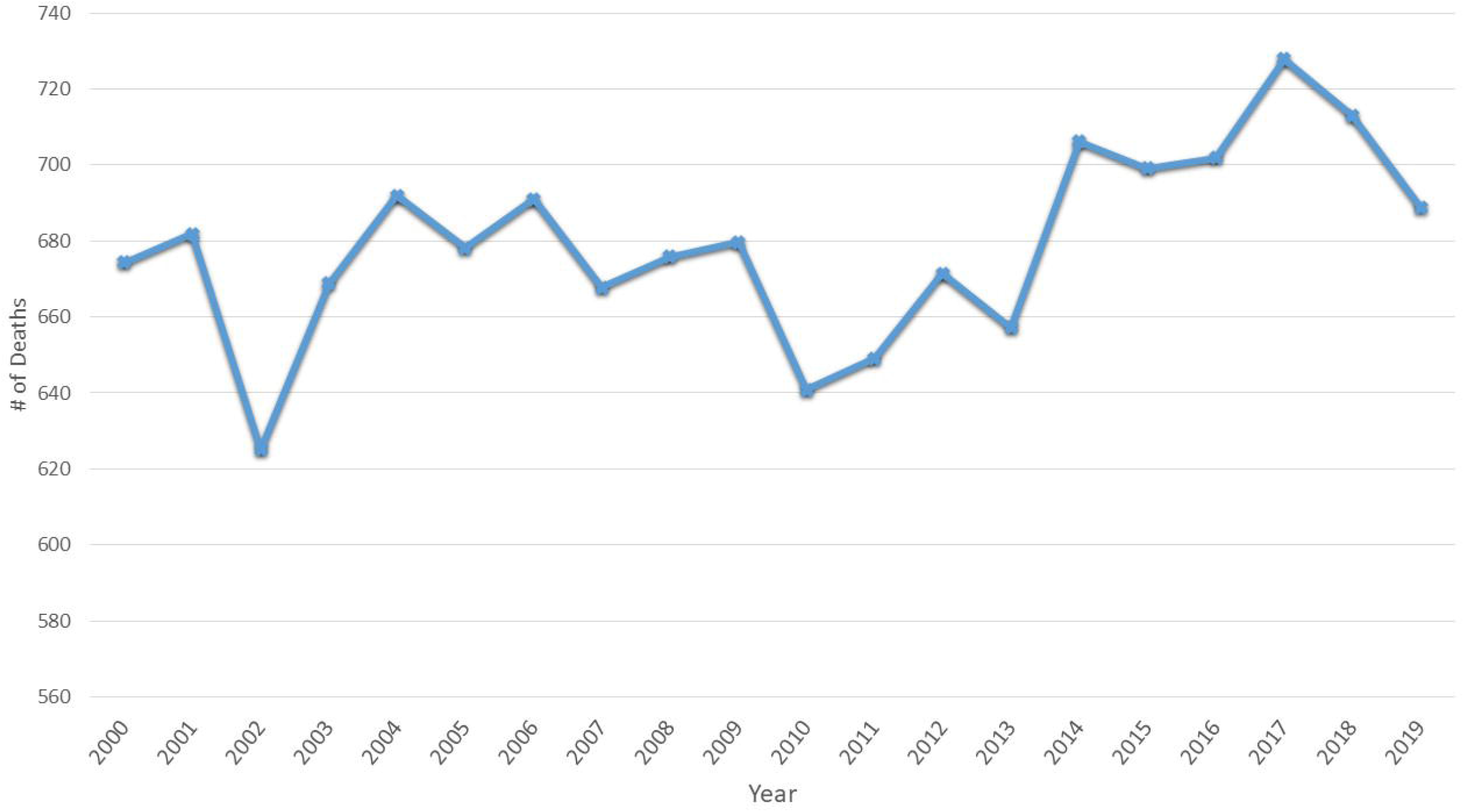
Number of drowning deaths in Zambia from, 2000 - 2019 (Source WHO Global Health Estimates)

Zambia’s diverse geographical landscape, encompassing over 15 major rivers (e.g., Zambezi, Kafue) and numerous lakes (e.g., Tanganyika, Bangweulu, Kariba), contributes to elevated drowning risks, particularly in rural communities reliant on these water bodies for transportation, fishing, and water collection (Zambia Statistics Agency, 2022). Swimming pools are scarce, especially in rural areas, increasing dependence on natural water bodies and amplifying risks (Tyler et al., 2017). Cultural practices, such as artisanal fishing and water fetching, expose individuals, particularly children, to drowning risks, compounded by socio-economic factors like poverty and limited safety infrastructure (Tyler et al., 2017; Miller et al., 2019).

Despite this alarming burden, drowning prevention has been overshadowed in public health priorities, even though cost-effective interventions—such as pool fencing, mandatory life jacket use, swimming and water safety education, and community-based supervision programs—can significantly reduce mortality (World Health Organization, 2014; Meddings et al., 2021). Rapid urbanization and limited access to swimming lessons disproportionately affect low-income communities, exacerbating vulnerabilities (Miller et al., 2019). Drowning rates in LMICs like Zambia are over three times higher than in high-income countries, driven by inadequate supervision, limited swimming education, and unsafe watercraft (World Health Organization, 2014). The United Nations General Assembly’s resolution on global drowning prevention (United Nations, 2021) and its endorsement at the World Health Assembly (World Health Organization, 2023) provide a framework for Zambia to strengthen localized strategies tailored to its environmental and cultural contexts.

Drowning prevention aligns with the International Health Regulations’ goal of addressing public health risks threatening global health security (World Health Organization, 2016). In Zambia, increasing flood disasters heighten drowning risks, particularly for children and water-based workers (Tin et al., 2024). Effective prevention requires multifaceted approaches, including physical barriers, community-based childcare, and school-based water-safety education (Meddings et al., 2021; World Health Organization, 2014). This study assesses Zambia’s drowning prevention landscape, examining demographic and environmental risk factors, existing policies, coordination mechanisms, and interventions through stakeholder perspectives. By identifying gaps and opportunities within Zambia’s cultural and resource context, it aims to inform sustainable interventions and serve as a model for Sub-Saharan Africa, contributing to global efforts to reduce preventable drowning deaths.

## Methodology

### Study Design

This study employed a convergent mixed-methods design to conduct a national-level situational assessment of drowning prevention in Zambia. Data were collected in 2022 using a WHO-structured questionnaire that captured quantitative data on drowning cases (e.g., Zambia Police statistics) and qualitative data through open-ended responses on risk factors, barriers, stakeholders, interventions, and opportunities. Individual questionnaire responses were consolidated into a single country submission through a stakeholder workshop, reflecting national consensus. Quantitative data were analyzed descriptively, while qualitative data underwent thematic analysis, with findings integrated during interpretation to assess convergence and divergence, ensuring a comprehensive assessment aligned with WHO’s global framework (Supplementary File 1).

### Participants and Sampling

Following the WHO Global Status Report methodology, purposive sampling ensured multisectoral representation to address the complex risk factors of drowning, including environmental hazards, policy gaps, and limited awareness, necessitating collaboration across sectors (Meddings et al., 2021; Jagnoor et al., 2021). The WHO questionnaire required 6–10 National Data Collaborators (NDCs) from diverse sectors. In Zambia, 10 participants were recruited: two government officials from health (surveillance/policy), two from disaster management (flood risk reduction), one from education (water safety curricula), one from maritime safety (boating regulations), two NGO representatives (community engagement), one community leader, and one public health expert. This exceeded the minimum requirement to ensure comprehensive sectoral coverage.

### Data Collection

Data collection utilizing a WHO-structured questionnaire administered in 2022. The questionnaire, designed specifically for the WHO Global Status Report on Drowning Prevention (2024), comprised approximately 30 questions across five domains: national perspectives (e.g., risk groups, locations), stakeholders and strategies, drowning data, interventions/legislation, and future opportunities. It included quantitative data on drowning cases (e.g., number, sex, age distribution from Zambia Police statistics), structured response questions (e.g., tick-box options on stakeholder coordination, intervention implementation), and qualitative open-ended responses (e.g., one-sentence summaries on risk factors, barriers, and opportunities) to reflect national consensus. This national data collection, led by the Ministry of Health for WHO (2024), required no ethical review as it was not human subjects research. Participants provided written consent via securely stored forms.

### Data Analysis

Qualitative data from open-ended questionnaire responses were analyzed inductively in Microsoft Excel and using Braun and Clarke’s thematic analysis framework (Tong et al., 2007). Two researchers independently coded responses, developing a coding tree with main themes (e.g., high-risk groups, coordination barriers) and subthemes (e.g., child vulnerability, data gaps) aligned with the questionnaire’s five domains. Data saturation was achieved after analyzing responses from eight participants, with two additional responses confirming no new themes. Quantitative drowning case data (participant-reported Zambia Police statistics) were analyzed using descriptive statistics (frequencies and percentages) to summarize deaths by sex and age group, with no additional statistical tests due to the limited dataset size. Findings were integrated during interpretation to identify convergence (e.g., male-dominated drowning deaths aligned with reported occupational risks) and divergence (e.g., stakeholder-reported child risks not reflected in age data), as per WHO’s convergent mixed-methods approach.

Data validation followed WHO’s methodology, with the stakeholder workshop serving as the primary mechanism to review and consolidate individual questionnaire responses into a single country submission. The Ministry of Health reviewed and verified the consolidated responses, checking for logical inconsistencies and alignment with national data sources (e.g., Zambia Police statistics). Supporting documents (e.g., legislation, policies) were reviewed for consistency with questionnaire responses, aligning with WHO’s validation process for country submissions.

### Results

This study presents findings from a WHO-structured questionnaire and stakeholder workshop conducted in 2022 with 10 stakeholders in Zambia’s drowning prevention efforts, contributing to a global drowning prevention assessment. The questionnaire, comprising approximately 30 questions, collected qualitative data through open-ended responses and quantitative data on drowning cases, consolidated into a single country submission. All 10 participants (2 health officials, 2 disaster management officials, 1 education official, 1 maritime safety official, 2 NGO representatives, 1 community leader, 1 public health expert) completed the questionnaire and stakeholder workshop. Findings are organized into five thematic subsections—National Perspectives, Stakeholders and Strategies, Drowning Data, Interventions and Legislation, Future Opportunities—derived from thematic analysis of qualitative questionnaire responses, integrated with the quantitative drowning case data where relevant. Table 1 summarizes the qualitative findings, highlighting key themes and representative stakeholder perspectives.

**Table 1:**
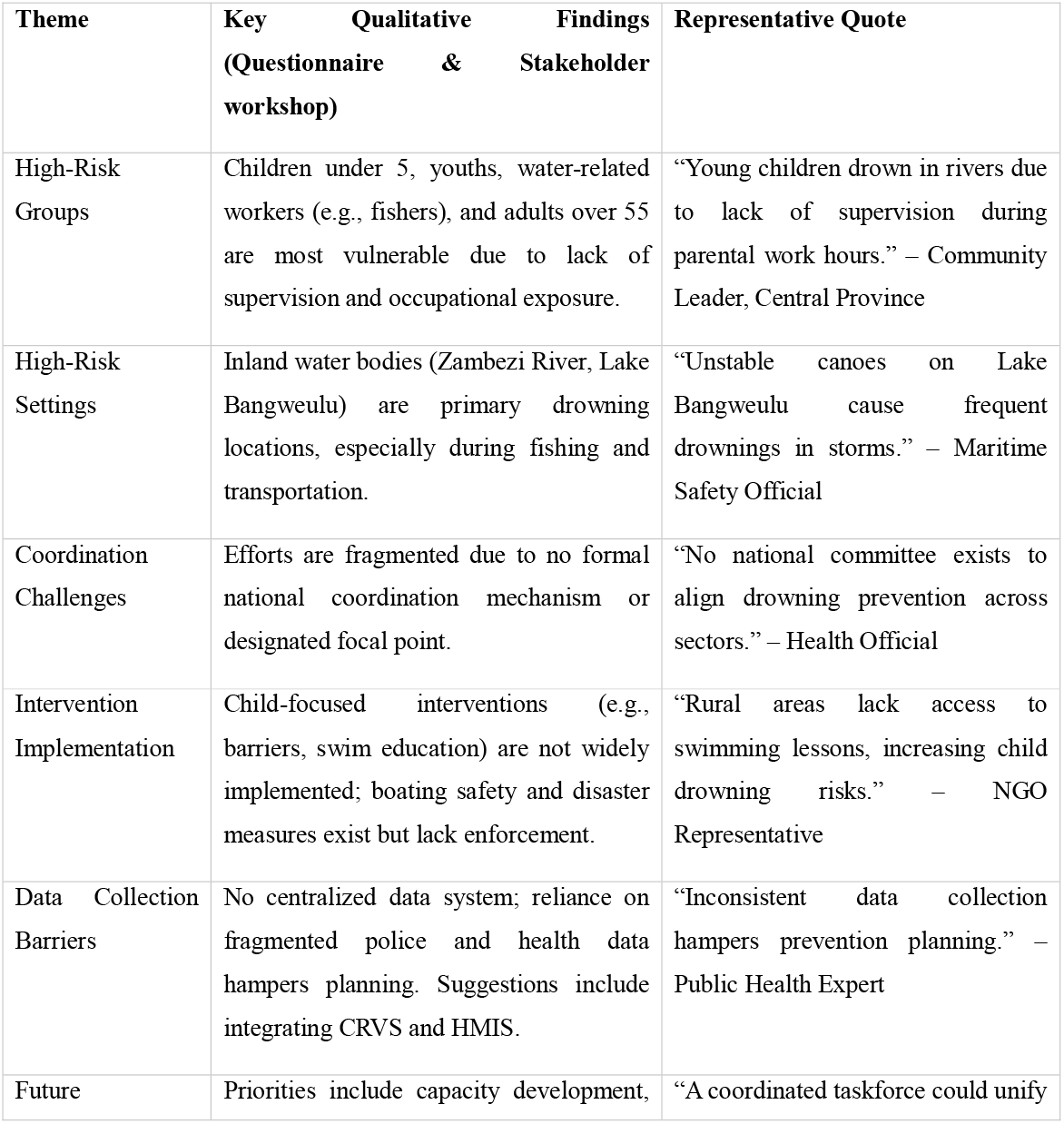
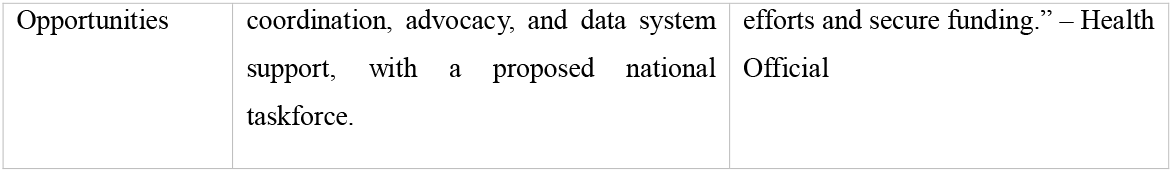
Summary of Qualitative Findings on Drowning Prevention in Zambia.

### National Perspectives on Drowning

Stakeholders reported that children under 5, youths, water-related workers such as fishers, and adults over 55 are the most vulnerable to drowning, based on questionnaire responses and stakeholder workshop. A community leader from Central Province stated, “Young children drown in rivers due to lack of supervision during parental work hours.” Inland water bodies, particularly the Zambezi River and Lake Bangweulu, were identified as high-risk settings, with a maritime safety official noting, “Unstable canoes on Lake Bangweulu cause frequent drownings in storms.” Drowning incidents were reported to occur primarily during water transportation, fishing, and recreational swimming, with daily activities like fetching water also posing risks. An education official highlighted flooding as a significant event increasing drowning risk, stating, “Floods in rural areas catch communities unprepared, leading to drownings.”

### Stakeholders and Strategies for Drowning Prevention

Questionnaire responses indicated that health, disaster management, and maritime safety sectors contribute to drowning prevention programs, but efforts are fragmented due to the absence of a formal national coordination mechanism. A health official stated, “No national committee exists to align drowning prevention across sectors.” Stakeholders reported a broader national strategy that includes drowning prevention, but it lacks specific, measurable targets or evaluation mechanisms. An NGO representative noted, “The national strategy mentions drowning but has no clear goals or monitoring systems.” Ongoing reviews of the Inland Water Shipping Act were highlighted as an opportunity to strengthen boating safety regulations, suggesting a pathway for improved policy implementation.

### Drowning Data

Participant-reported Zambia Police data from the questionnaire indicated 89 drowning deaths in 2022, with 88.8% male (n=79) and 11.2% female (n=10), primarily among individuals aged 30–49 years (n=50) (Figure 2). The age distribution included 5–14 years (n=20), 15–29 years (n=15), 50–69 years (n=4), with no deaths in 0–4 or 70+ years (Figure 3). This aligns with WHO Global Health Estimates (2019: 5.8 per 100,000; 2021: 4.9 per 100,000), though the lower police-reported figure suggests underreporting (World Health Organization, 2019, 2024). Stakeholder responses supported these data, with a public health expert stating, “Men’s occupational water exposure, like fishing, increases their drowning risk.” Stakeholders reported no centralized drowning data system, relying on fragmented sources such as police and health sector records. A disaster management official noted, “Inconsistent data collection hampers prevention planning.” Suggestions included integrating Civil Registration and Vital Statistics (CRVS) and Health Management Information Systems (HMIS) to enhance surveillance.

**Figure 2:**
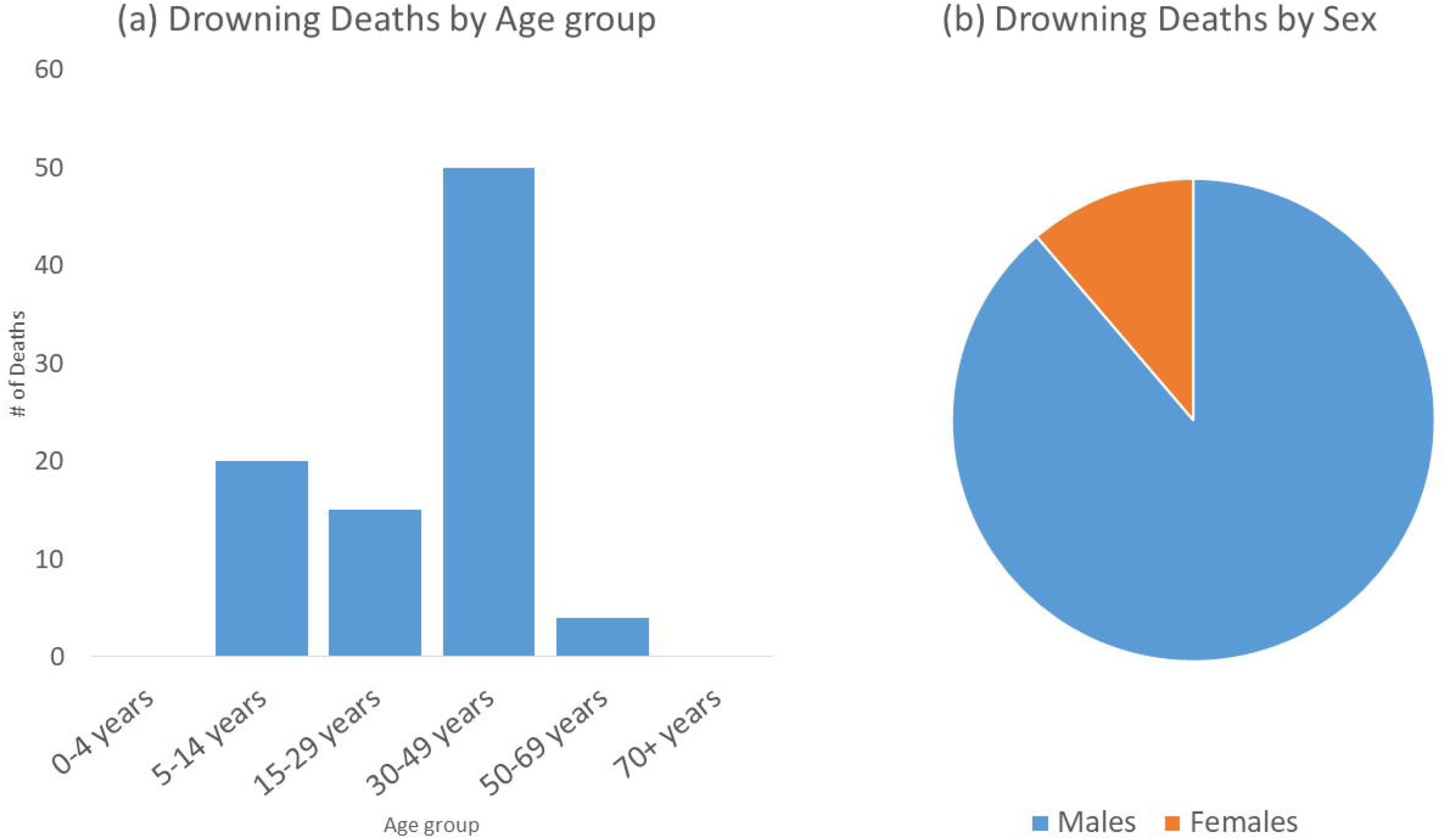
Drowning deaths in Zambia (2022) (Source: Zambia Police, participant-reported)

### Interventions and Legislation for Drowning Prevention

Stakeholders reported that child-focused interventions, such as physical barriers (e.g., well covers, pool fencing) and swim education programs, are not widely implemented, particularly in rural areas. An NGO representative stated, “Rural areas lack access to swimming lessons, increasing child drowning risks.” No laws mandate fencing around public or private pools, with a community leader noting, “Open wells and ponds pose risks to children without barriers.” Boating safety regulations and life jacket laws exist nationally, but enforcement is inconsistent. A maritime safety official said, “Life jacket use is low due to limited awareness and enforcement.” The Inland Water Shipping Act (Cap. 466) sets minimum safety standards for vessels, but stakeholders reported weak implementation. Disaster risk reduction interventions, such as flood warning systems, have national coverage but limited reach in remote areas, with a disaster management official stating, “Flood warnings rarely reach rural communities.” Search and rescue services were reported to have limited sub-national reach, constraining response capabilities.

### Future Opportunities for Drowning Prevention

Stakeholders prioritized capacity development, coordination, and advocacy for strengthening drowning prevention, based on questionnaire responses and a stakeholder workshop. A public health expert stated, “Training community lifesavers could reduce drowning deaths.” Technical support for improving data systems and sharing regional best practices was emphasized, with an NGO representative noting, “Learning from other countries’ successes could guide Zambia’s strategies.” Stakeholders proposed establishing a national drowning prevention taskforce, with a health official stating, “A coordinated taskforce could unify efforts and secure funding.” Public awareness campaigns were also highlighted, with a community leader noting, “Public campaigns could shift attitudes toward water safety.”

## Discussion

This study provides a comprehensive assessment of drowning prevention in Zambia, drawing on a consolidated WHO-structured questionnaire and stakeholder workshop with 10 stakeholders from diverse sectors (health, disaster management, education, maritime safety, NGOs, community leadership, and public health expertise). The findings highlight critical gaps in drowning prevention efforts, aligning with global evidence on drowning as a preventable public health issue (World Health Organization, 2019; Meddings et al., 2021). The qualitative themes—high-risk groups, high-risk settings, coordination challenges, intervention implementation barriers, data collection limitations, and future opportunities—converge with the quantitative drowning case data to underscore the urgent need for targeted interventions and systemic improvements in Zambia.

Stakeholders identified children under 5, youths, water-related workers (e.g., fishers), and adults over 55 as the most vulnerable to drowning, consistent with global patterns where children and occupationally exposed adults face elevated risks due to lack of supervision and hazardous water activities (World Health Organization, 2014; Jagnoor et al., 2021). The high-risk settings of inland water bodies, such as the Zambezi River and Lake Bangweulu, reflect Zambia’s geography, where reliance on rivers and lakes for transportation and livelihoods amplifies drowning risks, particularly during storms or flooding. These findings align with studies in other low- and middle-income countries (LMICs), where natural water bodies are primary drowning locations due to limited infrastructure and safety measures (Rahman et al., 2017). The reported risks during daily activities (e.g., fetching water) and flooding events highlight the intersection of environmental and socioeconomic factors, reinforcing the need for context-specific interventions tailored to Zambia’s rural and water-dependent communities.

The absence of a formal national coordination mechanism, as reported by stakeholders, mirrors challenges in other LMICs where fragmented efforts hinder drowning prevention (Franklin et al., 2020). The lack of a designated focal point and measurable targets within Zambia’s national strategy limits accountability and progress, a gap also noted in regional studies (e.g., Jagnoor et al., 2021). The ongoing review of the Inland Water Shipping Act offers a promising opportunity to strengthen boating safety regulations, aligning with WHO recommendations for legislative enforcement (World Health Organization, 2014). However, the reported fragmentation suggests that establishing a national drowning prevention taskforce, as proposed by stakeholders, could unify efforts and enhance resource allocation, a strategy successfully implemented in countries like Bangladesh (Rahman et al., 2017).

The Zambia Police data (89 deaths in 2022, 88.8% male, 11.2% female, primarily aged 30–49 years) indicate a male-dominated drowning burden, converging with qualitative reports of occupational water exposure among fishers and transport workers. This aligns with WHO Global Health Estimates (2019: 5.8 per 100,000; 2021: 4.9 per 100,000), though the lower police-reported figure and 2021 rate suggest underreporting and possible methodological adjustments (World Health Organization, 2019, 2024). Stakeholders identified youths, young adults, and water-related workers as the most vulnerable, consistent with 2022 data showing 50 deaths in the 30–49 age group (World Health Organization, 2024). The absence of recorded deaths in 0–4 and 70+ years contrasts with stakeholder perceptions of children under 5 and adults over 55 as high-risk, possibly reflecting underreporting or broader regional trends (Jagnoor et al., 2021). The high-risk settings of inland water bodies align with studies in other LMICs (Rahman et al., 2019). The absence of a formal national coordination mechanism mirrors challenges in other LMICs (Peden et al., 2020). Integrating Civil Registration and Vital Statistics (CRVS) and Health Management Information Systems (HMIS), as suggested by stakeholders, could enhance data accuracy and inform evidence-based interventions, a priority also highlighted in WHO guidelines (World Health Organization, 2024).

The limited implementation of child-focused interventions, such as physical barriers and swim education, particularly in rural areas, reflects resource constraints and infrastructure gaps common in LMICs (Franklin et al., 2020). The absence of pool fencing laws and weak enforcement of boating safety regulations, including life jacket use, align with findings from other African contexts where regulatory frameworks exist but lack practical application (Jagnoor et al., 2021). Similarly, the limited reach of flood warning systems and search and rescue services in remote areas underscores disparities in access, a challenge compounded by Zambia’s rural geography. These findings highlight the need for scalable, community-based interventions, such as training local lifesavers, as proposed by stakeholders, which has proven effective in similar settings (Rahman et al., 2017).

Stakeholders’ prioritization of capacity development, coordination, advocacy, and data system support aligns with WHO’s global drowning prevention framework (World Health Organization, 2024). The proposed national taskforce and public awareness campaigns could address coordination and awareness gaps, drawing on successful models from other LMICs (Franklin et al., 2020). Technical support for data systems and cross-country learning, as suggested, could leverage regional best practices, such as community swim programs in Bangladesh (Rahman et al., 2017). These opportunities emphasize the importance of multisectoral collaboration and WHO support to build sustainable drowning prevention strategies in Zambia.

## Limitations

This study has several limitations. The consolidation of questionnaire responses into a single country submission, while aligning with WHO protocols, may have obscured individual stakeholder variations, potentially limiting the granularity of findings. The sample of 10 participants, although meeting WHO’s requirement of 6–10 respondents across multiple sectors, may not fully capture the diversity of perspectives in Zambia’s complex drowning prevention landscape. The quantitative data were limited to Zambia Police statistics (89 deaths), which likely underreport the true drowning burden compared to WHO estimates (600 deaths annually), reflecting data collection challenges. Logistical constraints prevented returning individual questionnaire responses to participants, though the stakeholder workshop ensured consensus. Finally, the study’s focus on stakeholder perceptions may not fully reflect on-the-ground realities, necessitating further community-level research.

## Recommendations

To address the identified gaps, we recommend establishing a national drowning prevention taskforce to coordinate multisectoral efforts, set measurable targets, and secure funding, as suggested by stakeholders. Community-based interventions, such as swim education and physical barriers around water bodies, should be prioritized, particularly in rural areas near the Zambezi River and Lake Bangweulu. Strengthening the Inland Water Shipping Act’s enforcement, including mandatory life jacket use, could reduce occupational drownings. Integrating CRVS and HMIS into a centralized drowning data system would enhance surveillance and inform policy. Public awareness campaigns, tailored to local contexts, could shift attitudes toward water safety, while WHO-supported capacity development and cross-country learning could accelerate progress. Future research should engage community members to validate stakeholder perceptions and assess intervention feasibility.

## Conclusion

This study highlights Zambia’s urgent need for enhanced drowning prevention, driven by a male-dominated drowning burden (89 deaths in 2022, 88.8% male, primarily aged 30–49 years) linked to occupational exposure among fishers and transport workers in inland water bodies. Systemic gaps— no national coordination mechanism, fragmented data systems, weak enforcement of boating regulations, and limited rural interventions (e.g., swim education, barriers)—exacerbate the burden, aligning with challenges in other low- and middle-income countries (World Health Organization, 2024). The absence of lifeguard or pool fencing legislation further limits prevention efforts. Recommendations include establishing a national drowning prevention taskforce to unify multisectoral efforts (health, disaster management, maritime safety, and additional sectors like fire and agriculture), strengthening the Inland Water Shipping Act for mandatory life jacket use, and integrating Civil Registration and Vital Statistics (CRVS) and Health Management Information Systems (HMIS) for robust surveillance. Community-based interventions, such as swim education and physical barriers, and public awareness campaigns tailored to rural contexts can address high-risk groups (youths, young adults, water-related workers). By leveraging WHO technical support and regional best practices, Zambia can build sustainable strategies to reduce drowning mortality, contributing to global health security and serving as a model for Sub-Saharan Africa.

## Acknowledgments

The authors would like to express their gratitude to the Zambian Ministry of Health, the Disaster Management and Mitigation Unit, The Ministry of Transport, the Olympic Youth Development Centre (OYDC), the Zambia Police Service and the participating non-governmental organizations for their valuable contributions and support throughout this study. We also acknowledge the assistance provided by our research team members in data collection and analysis.

## Declaration

The opinions, findings, and conclusions expressed in this paper are those of the authors and do not necessarily represent the official views, decisions, or policies of the World Health Organization, the Ministry of Health, or the Zambia National Public Health Institute.

## Data Availability

The datasets generated and/or analyzed during the current study are available from the corresponding author on reasonable request.

